# Reclaiming health: a qualitative, explorative study of long covid recovery journeys involving mind-body approaches

**DOI:** 10.64898/2026.02.21.26345052

**Authors:** Christine Deurman, Vera Brinkman, Nienke Slagboom, Jet Bussemaker, Hedwig Vos

## Abstract

**Objective:** This study explored the recovery experiences of individuals who report having (largely) recovered from long covid and who attributed their improvement to mind-body approaches.

**Design, setting and participants:** We conducted an explorative qualitative study using purposive recruitment through social media and snowball sampling. Eighteen adult women (aged 37-62 years), who self-identified as having had long covid and having substantially recovered through mind-body approaches participated in semi-structured interviews. Data were analysed using Saunders’ practical thematic analysis.

**Results:** Despite variation in personal narratives, a common trajectory emerged: participants moved away from a biomedical explanatory model towards one centred on nervous system dysregulation. This shift, sometimes following initial scepticism, was often described as a turning point, sparking hope and motivation to engage in self-directed strategies. Recovery was not linear but an iterative process, involving cycles of practice, reflection (especially when progress stagnated) and adaptation of mind-body techniques. Over time, participants gained insights into contributing factors and, in many cases, made intentional life changes to support ongoing recovery. These patterns echo findings from previous research on mind-body approaches in chronic pain and chronic fatigue, and align with neuroscientific perspectives on symptom generation.

Most participants navigated this process without formal clinical support, relying instead on online communities and actively avoiding sources of (biomedical) information that conflicted with their new understanding.

**Conclusions:** While causal inferences cannot be drawn from qualitative data, this study highlights potential mechanisms that may underpin recovery for people with long covid using mind-body approaches. Further research is needed to develop structured interventions, and to evaluate their efficacy and safety.

Future research should also explore how prevailing narratives within healthcare and society influence treatment engagement and recovery trajectories.

**STRENGTHS AND LIMITATIONS OF THIS STUDY:**

- This is the first study exploring experiences of recovery from long covid using mind-body approaches.
- In-depth, real-world accounts capture the lived-experiences over time and allow in-depth exploration if the recovery process, while the semi-structured design facilitates the emergence of themes rarely captured in clinical research.
- Generalisability is limited due to self-identified long covid status, lack of formal diagnostic verification, absence of strict definitions of ‘mind-body approaches’ and ‘recovery’, and a relatively homogenous sample (mostly highly educated women).

## INTRODUCTION

Shortly after the onset of the covid-19 pandemic, case reports began to emerge describing individuals who experienced a wide range of persistent symptoms following SARS-CoV-2 infection [1–4]. This condition is now commonly referred to as post covid-19 condition, or long covid.

Despite substantial research into its epidemiology and pathophysiology (such as [5–8]), the underlying mechanisms of long covid remain poorly understood - particularly in cases without clear biomedical correlates following mild or moderate infection. Currently available pharmaceutical treatments offer, at best, moderate relief [9], and spontaneous recovery of long-lasting severe symptoms appears unlikely [10–13].

From the early stages of the pandemic, healthcare professionals and patients alike observed similarities between long covid symptoms and symptoms seen in other chronic conditions, such as myalgic encephalopathy/chronic fatigue syndrome (ME/CFS) and persistent physical symptoms (PPS). This prompted consideration of alternative perspectives on the origin and management of long covid symptoms, at least for a subset of patients. Within this alternative framework, symptoms are not solely attributed to simple, linear, pathophysiological relationships but are seen as the result of complex and dynamic interactions among biological, psychological, and social factors - drawing on established knowledge from the field of functional somatic symptoms [14–16].

Concurrently, numerous accounts began to appear on social media of individuals reporting recovery from long covid while using what they referred to as mind-body approaches, among others at websites of the Emovere Foundation, Sirpa UK and in the Dutch Facebook group “*Mind body syndroom en long Covid NL*” (hereafter referred to as “the Dutch mind-body Facebook group”).

While the term ‘mind-body’ lacks a universally agreed definition, it generally refers to interventions that assume a significant role of brain-body interactions and behaviour in the perception and persistence of symptoms. These approaches often involve strategies intended to influence these interactions through knowledge, emotional processing, and behavioural change [17–19].

We recognise that the use of mind-body techniques in long covid is a contentious issue. As similarities between long covid, other post-infectious syndromes and PPS have become more apparent, debates about the nature and etiology of the condition have intensified. These discussions echo historical experiences of people with ME/CFS, many of whom have felt and continue to feel dismissed. This pattern reflects a broader issue of epistemic injustice, where patients’ lived experiences and knowledge have been systematically undervalued in clinical and research contexts [20–23].

As a result, current discourse largely prioritises biomedical research and pharmacological solutions, and mind-body work may be perceived as challenging established biomedical frameworks. However, given the scale of long covid and the paucity of (highly) effective medicines for long covid, it is imperative to study these recovery stories. They may offer valuable insights for individuals navigating similar conditions, for healthcare professionals seeking to support them and for researchers who are developing interventions.

A few qualitative studies have explored the lived experiences of recoverees who applied mind-body approaches for chronic back pain and for ME/CFS [24–26]. However, to our knowledge little is known about the lived experiences of individuals who have used mind-body approaches in long covid. This qualitative study therefore aims to explore the recovery experiences of individuals who report having (largely) recovered from long covid and who attribute their improvement to approaches they identify as mind-body. By examining their retrospective reflections on the recovery process, this study seeks to provide insight into the common key elements and patterns of these experiences, and to inform future research and care strategies for individuals with long covid.

## METHOD

### Study design and study setting

This exploratory study involved semi-structured interviews with 18 women who have (largely) recovered from long covid. Participants were asked to retrospectively reflect on their recovery process.

### Patient and public involvement

This study was conducted as part of the ‘Emovere project’ on chronic pain and PPS. This citizen science project was initiated by a group of recovered patients and health care professionals committed to raising awareness about the mind-body connection and the potential role of suppressed emotions in PPS (the non-profit Emovere Foundation). The citizen science project was supported by a consortium including the Emovere Foundation, general practitioners, health insurers, third sector organisations and Leiden University Medical Center (LUMC).

For the current study, a patient representative from the Emovere Foundation was asked for advice on recruitment of participants. The administrator of the Dutch mind-body Facebook group, a recovered long covid patient herself, supported participant recruitment by allowing a call for recruitment to be posted online.

### Researcher characteristics and positionality

The research team consisted of five female researchers with differing professional backgrounds, personal experiences and relationships to the study topic. CD and HV are both clinicians, one with a focus on public health, the other on general practice. CD’s years of interest in PPS, patients’ recovery stories and her recent training in mind-body therapies complemented HV’s primary care experiences and approach. VB entered the project as a medical student unfamiliar with chronic symptoms or mind-body approaches, initially viewing them sceptically, which added a fresh lens. NS contributed an anthropological perspective, emphasizing systemic context, and JB brought a policy-oriented view on patient involvement and impact on health system level.

None of the researchers had any prior relationship with the study participants, nor any personal experience with long covid or ME/CFS. Throughout the research process, the team engaged in regular reflexive conversations to acknowledge and mitigate potential bias stemming from their backgrounds, beliefs and roles.

### Recruitment and sampling

Potential participants were recruited in March and April 2024 through the country wide network of the Emovere Foundation and the Dutch mind-body Facebook group (with over 3,400 members at the time of recruitment). A purposeful, criterion-based sampling strategy was employed, an approach informed by a similar study on recovery in ME/CFS [25]. Inclusion criteria were: age ≥18 at the time of symptom onset, self-reported symptoms persisting for at least three months following the presumed initial SARS-CoV-2 infection, and the experience of either full recovery or substantial progress towards recovery. A positive SARS-CoV2 test was not required. We included participants who attributed their recovery to what they themselves referred to as a ‘mind-body approach’. The term mind-body approach was intentionally left undefined in order to include a range of techniques and practices that participants considered to fall within this category.

Over 40 individuals responded to our recruitment posts. As this was an exploratory study, we could not anticipate the degree of variation in recovery experiences. Therefore, we started with contacting 20 respondents, of whom 18 met the eligibility criteria and agreed to participate in interviews, conducted between April and July 2024. Following the provision of detailed study information, both written and verbal informed consent were obtained from all participants prior to the interviews. Data saturation was reached after 16 interviews; however, we completed all 18 as planned. The remaining respondents were thanked for their interest.

Table 1 describes characteristics of the participants of the study, including their self-reported symptoms. Table 2 describes the estimated moment in time of their presumed infection, based on the timeline of events that was created with the participants. Other spontaneously reported symptoms can be found in the supplemental material.

**Table 1.** Sociodemographic and other characteristics of the participants of the study.

|  |  |
| --- | --- |
| Age (yrs), mean (range) | 49.7 (32-67) |
| Age (yrs), n (%) |  |
| 30-39 | 2 (11.1) |
| 40-49 | 5 (27.8) |
| 50-59 | 9 (50) |
| 60-69 | 2 (11.1) |
| Sex, n (%) |  |
| Female | 18 (100) |
| Education level*, n (%) |  |
| lower education level | 0 (0) |
| intermediate education level | 5 (27.8) |
| higher education level | 13 (72.2) |
| Religious affiliation, n (%) |  |
| Christian | 2 (10.5) |
| None | 16 (89.5) |
| Born in the Netherlands, n (%) |  |
| Yes | 17 (94.7) |
| No | 1 (5.3) |
| Marital status, n (%) |  |
| Single | 1 (5.6) |
| living together or married | 15 (72.2) |
| Divorced | 2 (11.1) |
| Number of self-reported symptoms, average (range) | 11.7 (5-17) |
| Top 5, n |  |
| Hypersensitivity to light and/or sound | 17 |
| Fatigue | 16 |
| Difficulties to concentrate (“brain fog”) | 16 |
| Shortness of breath | 15 |
| Palpitations | 14 |
| Other self-reported symptoms (n):<br>Sleeping problems (13), memory problems (13), low or irritable mood (12), headache (12), muscle ache (12), dizziness (12), changes in temperature (9), joint aches (9), (night)sweats (8), visual problems (8), chest pain (6), abdominal problems (4), loss of smell (4), cough (4), skin problems (4), sore throat (2). |  |
| * Lower educational level: Primary education and lower secondary education (vocational and general tracks). Intermediate educational level: Upper secondary education and intermediate vocational education. Higher educational level: Higher professional and academic education. |  |

**Table 2.** Estimated moment of infection.

| Estimated quarter of SARS-CoV-2-infection, n (%) | 2020 | 2021 | 2022 |
| --- | --- | --- | --- |
| Q1: January - March | 1 (5,6%) | 4 (22,2%) | 2 (11,1%) |
| Q2: April – June |  | 2 (11,1%) |  |
| Q3: July – September |  | 1 (5,5%) | 1 (5,6%) |
| Q4: October - December | 5 (27,8%) |  | 2 (11,1%) |

### Data collection and processing

Data were collected through a questionnaire about symptoms that was sent by email before the interview, and through semi-structured interviews. The questionnaire (supplementary material) was based on the top 20 of long covid symptoms [27] and was used to get an impression of the array of symptoms the participants had suffered from. Interviews took place either online or face-to-face, depending on participants’ preference, and were scheduled at a time and location convenient to each participant.

The interview guide was based on topics from existing qualitative studies on recovery from ME/CFS and chronic back pain [24, 25]. Topics included a summary of the illness trajectory before encountering mind-body work, how participants learned about mind-body concepts and how this affected their ideas about their illness and their therapeutic approach (see supplementary material).

CD conducted 16 interviews and a psychology master student conducted the last 2. Following the first two pilot interviews, minor revisions were made to the interview guide based on feedback from the research team. During each interview, fieldnotes were taken and the interviewer drew a visual timeline of symptoms and events with the participant to aid memory, clarify the recovery trajectory and triangulate the data, as used in previous studies [28, 29].

The interviews lasted between 40 and 80 minutes, with one exception of 140 minutes, which reflected a particularly long and complex recovery journey. Following participant consent, face-to-face interviews (n=6) were audio-recorded and online interviews (n=12) were additionally video-recorded. Interviews were transcribed verbatim using Amberscript, a secure transcription service. CD reviewed all transcripts for accuracy, removed identifying information, and pseudonymised by assigning aliases to maintain participant confidentiality, while supporting narrative coherence and interpretive depth in the qualitative analysis.

### Data analysis

Data analysis was conducted by CD and VB under the supervision of the research team (HV, NS). A practical inductive approach to thematic analysis was employed [30], consisting of reading, coding and theming. CD and VB began by familiarising themselves with the interview data and taking initial notes. Four transcripts were then coded independently using open coding. Transcripts were read and coded line by line in Microsoft Word, and a coding tree with corresponding quotes was organised in Microsoft Excel.

Following the initial phase of open coding, CD and VB met to compare and consolidate their codes, reviewing the supporting quotations to ensure consistency and removing duplicate codes, with support from HV. This process resulted in a preliminary codebook, which was then applied to two additional transcripts. New codes that emerged during this phase were added (after consolidation) to the codebook, which was subsequently used by VB to code the remaining transcripts.

To identify overarching themes, two in-depth sessions were held with CD, VB, and HV. Themes were developed based on the research question, and included common elements in narratives and recurrent patterns in the participants’ recovery trajectories. This was done manually, by grouping printed codes on a very large table. Axial coding was subsequently done in Atlas.ti v24.

## FINDINGS

In this qualitative study we explored the recovery experiences of individuals who report substantial improvement from long covid, attributed to mind-body approaches, and we identified common elements and patterns in their retrospective accounts.

Table 3 provides background of the participants: their age, their social situation around the time they fell ill and their reported illness-related impairment.

**Table 3.**
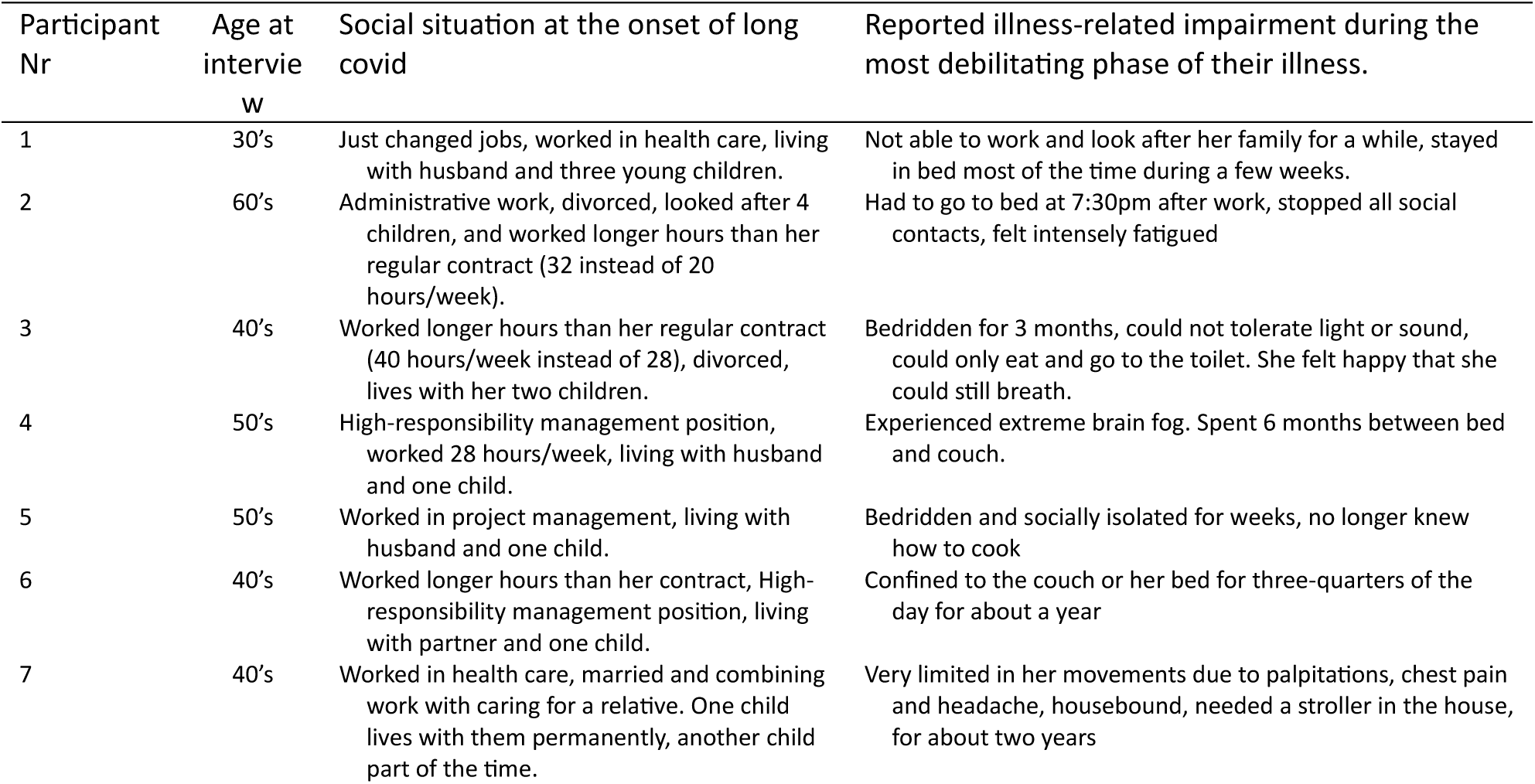
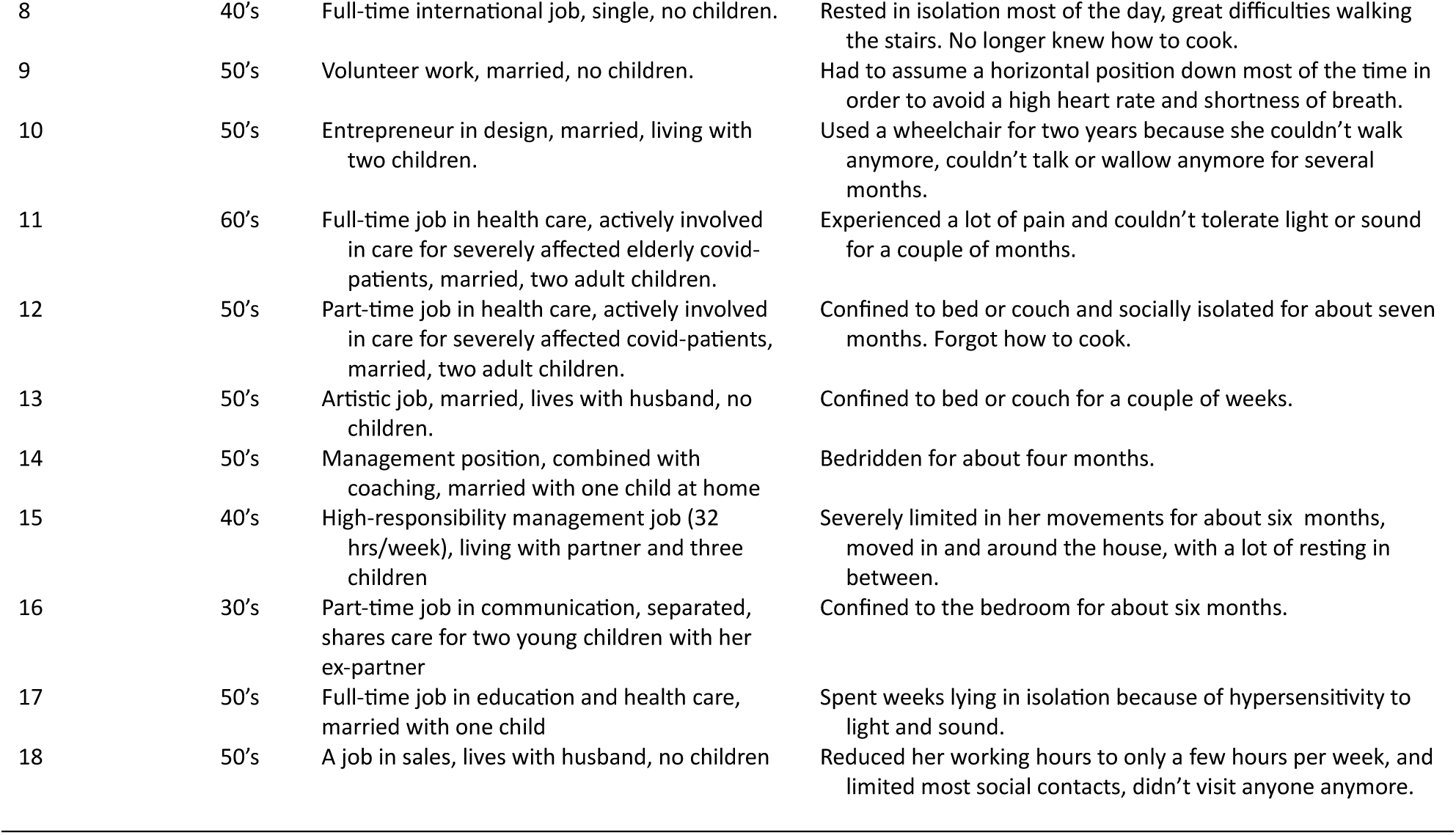
Overview of participants with background information and illness related information.

Participants had often already experienced a long and distressing search for explanations and treatments that had brought insufficient relief. Before starting mind-body approaches, participants had received an average of 2.4 treatments within mainstream healthcare (range 0-4), and two types of treatment within complementary healthcare (range 0-4). Physiotherapy and occupational therapy were the most commonly reported treatments within mainstream healthcare, whereas acupuncture and orthomolecular therapy were most frequently reported among complementary and alternative medicine (supplemental material). The duration of illness before engaging with a mind-body approach intervention varied widely, ranging from 5 to 32 months (median 23 months). As illustrated in table 2, many described long covid as an isolating and disruptive experience, marked by debilitating symptoms and repeated disappointments with mainstream healthcare.

At the time of the interviews, patients described themselves as (largely) recovered. Figure 1 illustrates common elements and themes in participants’ description of their recovery journey, from the initial encounter to an iterative cycle of starting to practice mind-body approaches while learning more about these techniques, noticing changes and reflecting on factors that play a role in their illness and recovery.

**Figure 1.**
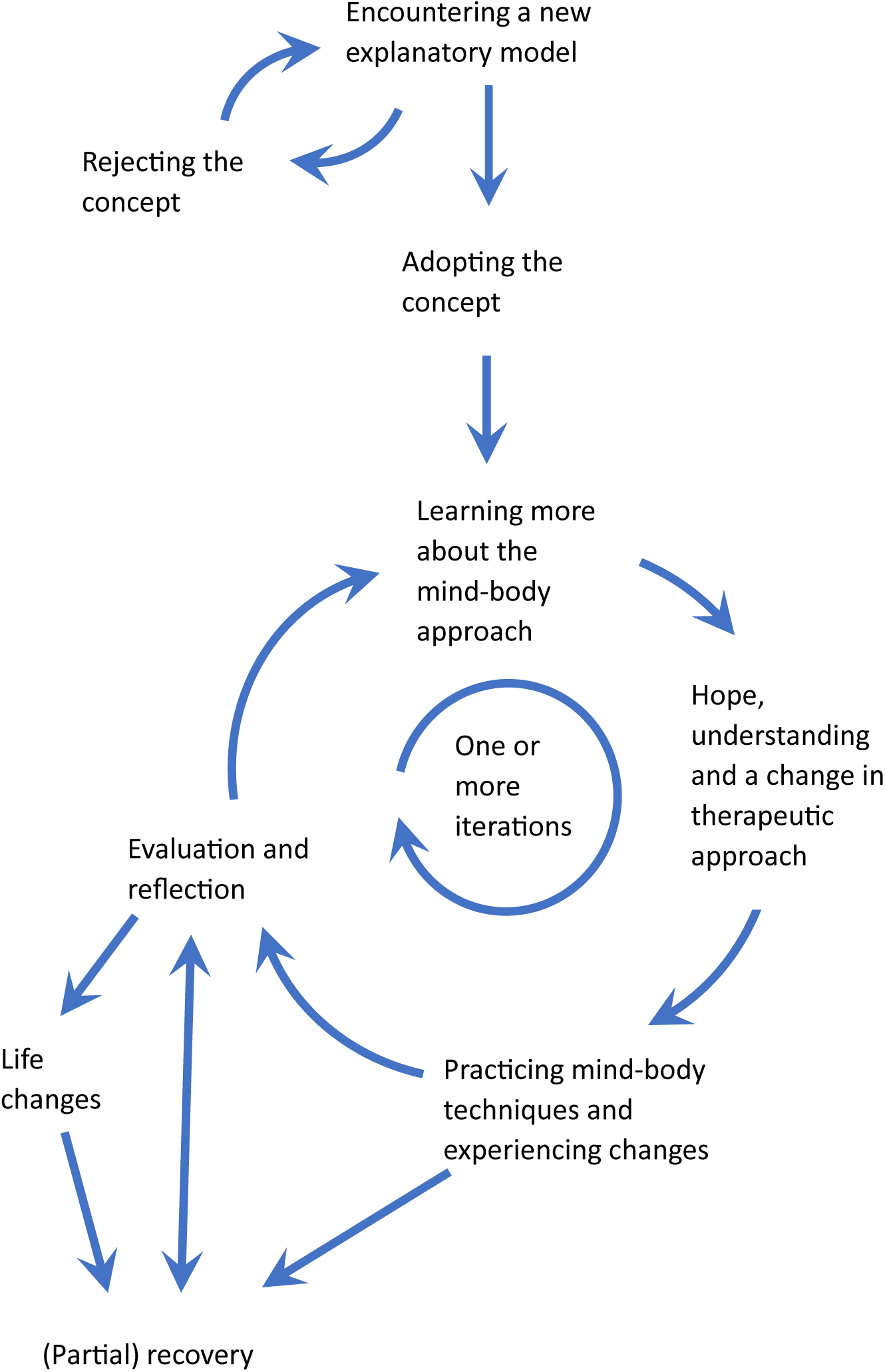
Themes and elements in participants’ recovery journeys

### Encountering a new explanatory model for long covid symptoms

> “I started reading a book about long covid. And it was just one moment of recognition after another. I had so many symptoms that I honestly thought: someone must have been spying on me and written a book about it. Everything in it just made perfect sense” (Karen)

A lack of noticeable improvement in symptoms, despite the treatments they had received, prompted participants to seek alternative explanations for their suffering from long covid symptoms. They consulted a wide range of sources, including social media platforms, websites, books, and podcasts (see supplemental material). In these sources, they found narratives of recovery or persistent symptoms shared by other patients who used mind-body approaches, shaping participants’ understanding and expectations. In one case an e-mail message from a health professional was the first introduction to a mind-body approach, and in another case an acquaintance spontaneously offered to mail some information.

The quote above is one of many that describe an immediate feeling of recognition. However, two distinct types of responses were observed following participants’ initial exposure to mind-body approaches: immediate adoption or initial rejection of the mind-body concept that their brain was an important driver of their long covid symptoms. Immediate adoption of this concept occurred when information about the mind-body approach altogether ‘just made sense’. In those cases, participants recognized both their symptoms and the new explanations for their suffering in this information.

Similar to Karen, Susan immediately accepted that her nervous system acted as an alarm that does not switch off: “*As if everyone inside your head is shouting: ‘We need to warn this woman, something’s wrong!’ That made perfect sense to me and [..] this explains what I’ve been feeling”*. The role of brain and nervous system was described by participants in terms like ‘dysregulation and dysautonomia’, the brain ‘being stuck’: ‘in fight-flight mode’, ‘in continuous stress-mode’, ‘on high alert’, ‘in survival-mode’, or ‘conditioned to sense danger (even when there’s none)’, often due to previous stressful times or traumas.

Initial rejection, the second response, emerged among those who felt the abovementioned explanations did not align with their personal experiences, lacked scientific credibility, or simply ‘sounded weird’. Elizabeth, for example, describes her initial rejection as follows: *“I didn’t recognise myself in stories about trauma or a difficult childhood, and I wasn’t in pain either. So for me, it didn’t click - my brain just didn’t believe it.”* And Iris was intrigued by the stories, but missed scientific evidence and therefore decided*: “I’m going to leave this alone, because it doesn’t feel solid enough to me.”*

Yet, over time - and often through repeated exposure and/or growing desperation - all participants became open to considering the new explanation for causes of their long covid symptoms: moving from a belief that their symptoms stemmed from (dangerous) physical damage, as a result of their primary SARS-CoV-2 infection, to viewing their suffering and symptoms as the result of a (largely) reversible mind-body interaction. This new explanation was adopted alongside the experienced physical reality of their symptoms: “*My symptoms were one hundred percent real, but simply generated by my brain*” (Elizabeth). And Helen used a metaphor to describe how the reality of her symptoms could align with the mind-body explanation: *“The symptoms are real, it’s just that my brain thinks I’m facing a lion or a tiger when I walk up the stairs.“*

### Learning more about the mind-body approach

> “I really dove into it (…). I thought: what kind of world is this? That’s when I came across all these different programmes. Thousands of people who had fully recovered. So much information. I felt like a complete beginner.” (Iris)

After the adoption of the mind-body explanation of long covid symptoms, participants wanted to learn more about the principles of mind-body approaches and how to implement these in their daily life. When searching for reliable information all but one participants faced several hurdles as a result of their illness: they were limited in their abilities to read and take in information, due to symptoms like brain fog, concentration difficulties and fatigue, or as Karen put it: “*I couldn’t process it (…), I just couldn’t take it in.”* And Susan mentioned that it took *“an absurd amount of effort”* due to her low energy level.

As a result of their limitations, participants had to come up with various strategies to gather the information they felt was necessary or helpful, such as listening to podcasts, audiobooks or YouTube instead of reading, or dividing the information in small bite-size portions, which was a time-consuming process.

Participants found workarounds for reading long texts: by listening to (Dutch) audiobooks, like “The way out” [31] and “Weg van de pijn” (Away from the pain) [32] or focusing on short messages on Facebook and YouTube. In addition, participants described having to navigate an overwhelming quantity of mind-body content available on- and offline, as illustrated above by Iris, predominantly in English, which often complicated finding information that fit their own situation. Several participants mentioned reading the Dutch summary of mind-body work they found in the Dutch mind-body Facebook group.

Despite the challenges described by participants, the information they were able to access marked the beginning of change because they developed a new perspective on their symptoms. And this in turn brought a renewed sense of hope and self-efficacy, and a new perspective on therapeutic approaches.

### Hope, a change in therapeutic approach and acceptance

> “It started for me with knowing [from recovery stories] that I could get better. Once that penny dropped – that I wasn’t broken, but that somehow my system had become misaligned – I could start looking for ways to set things straight again.” (Petra)

Petra illustrates a typical pattern, starting with the realisation that her physical symptoms may not stem from irreversible physical damage, which resulted in a new perspective on recovery. As participants learned more about mind-body approaches, three themes emerged. First, many reported a renewed sense of hope. Second, attributing their symptoms to a different underlying cause resulted in considering different treatment strategies. And third, a substantial part of the participants described that the first step of their new approach involved accepting the situation as it was.

#### Hope

Hope was derived from recovery stories, even when the described symptoms were different. Elizabeth for instance did not identify directly with the experiences shared in Nichole Sachs’ podcast [33], which predominantly focuses on chronic pain, but still found the stories *“really comforting - because everyone came out the other side, even after the most awful experiences. It was just nice to hear something so positive.”* And for Nicola, learning that people actually could recover was an eye-opener, providing a last resort: *“I thought: wow! (…) This is my last lifeline.”*

In addition, an underlying plausible rationale provided participants with new hope for recovery, and from the suggestion that they could influence their own recovery, as Petra illustrates: *“once I understood what was going on, and that it could pass, I thought why wouldn’t I be able to achieve that?”.* Ruth expressed a similar sentiment, but realised that it would take effort: *”on the one hand you become hopeful, on the other hand you have to start practicing yourself”*.

#### A change in therapeutic approach

Looking at the symptoms with a different view implied that a different treatment approach was needed. Petra, for instance, mentioned that a *“software problem”* needs *“reprogramming”*. Karen, who started noticing patterns in her symptoms, came to the conclusion that *“something isn’t being regulated properly - so I need to do something in my mind to help restore that regulation.”* The different treatment approach according to the new explanatory model focused on the connection between mind and body and on techniques that influence this connection.

This new approach did not solely involve the active adoption of mind-body practices; it also frequently entailed discontinuing previous therapies - most commonly physiotherapy and occupational therapy, but also various complementary or alternative treatments. Julie, for instance, realised she “*had to make a shift now*” - a decisive break from previous approaches in order to fully commit to working within the mind-body framework. And Angela describes her decision to stop previous therapies: *“I need to teach my body that it’s okay again - I need to reassure it. So I stopped physiotherapy, because I felt like I was constantly fighting and pushing myself. I stopped with all of it.”*

Participants began to avoid information they perceived as conflicting with their newly adopted outlook. Notably, many chose to disengage from other Facebook groups where symptoms were a frequent topic of discussion and stopped searching online for biomedical explanations of long covid. Some stopped using smartwatches for biometric monitoring. Helen illustrated: *“I stopped pacing, took off my smartwatch, left the Facebook group, and stopped looking things up altogether”*. These sources of information tended to emphasise symptom severity and biomedical causes of their symptoms, and were perceived to increase symptoms and anxiety.

#### Acceptance

Finally, participants underscored the significance of accepting their present circumstances as a vital step towards progress, in order to reduce system arousal. In this context, acceptance did not imply surrendering to a bleak prognosis, but rather a shift away from thinking in terms of ‘being broken’, needing a biomedical solution, towards a more present-centred way of living. Frances described why acceptance was important for her: *“If you don’t accept how things are now, you can’t begin with mind-body practices”*.

And acceptance of the current situation, in combination with a new found hope that recovery could be possible in the future, had direct impact on Deborah and Angela:

> “It gave me a sense of relief, I think - a kind of calming feeling, like: okay, we’re going to get through this. I really am safe. It’s going to be alright - not right away, but eventually.” (Deborah)

> “So stop fighting [the symptoms] and give it time - that became a kind of magic phrase for me. I thought: I won’t stay like this forever, I just need time, and that really helped me. Simply acknowledging that healing takes time.”(Angela)

### Experiential learning to apply mind-body techniques

> “I started reading that book (‘Weg van de pijn’ by Saskia de Bruin [32]) and began getting up every day, telling myself: I’m not ill, there’s nothing wrong, I no longer have covid. What I’m experiencing is tension and repressed emotions. That’s why I don’t feel well. I kept repeating that to myself like a kind of mantra. And whenever I thought something might be too much for me, I’d say: no, of course it’s not too much - because I’m not ill. So I’d just carry that suitcase upstairs, and day by day I had more energy. That really encouraged me. I thought: yes, this is it.” (Paula)

This quote by Paula typically illustrates common elements once participants started practicing a mind-body approach: the new perspective on their illness, the techniques applied, the ongoing effort it took, and changes they experienced when applying techniques.

Participants engaged with mind-body techniques differently: some focused on a single method or source, while others experimented with multiple mind-body techniques. In each of their descriptions, participants spoke of learning through experience, seeking what resonated with them personally or yielded tangible improvements. A list of the techniques they used can be found in the supplemental material.

The most frequently mentioned practices were 1. self-reassurance (by talking themselves), combined with the gradual resumption and build-up of activities, and 2. reframing the condition, symptoms, bodily sensations, and the recovery process using different terminology.

#### Self-assurance, combined with graded exposure

Self-reassurance was often applied during the activities that provoked symptoms such as walking, exposure to sunlight, or being in a noisy environment. Nicola, for example, used to feel during walking as if it was *“code red”* in her body, with sensations of *“muscle acidosis”* and her *“nerve pathways being on fire“*. While on a walk, she started telling herself about the benefits of walking, and after ten minutes, suddenly she *“snapped out of [these sensations]”.* She then decided to use the technique in other situations as well. For example, she used to experience severe brain fog in busy environments. So she started actively seeking out moments of exposure to many stimuli, to have *“corrective experiences”*: *“Because then your system learns that it **is** possible. That a shutdown doesn’t have to be the only option”*.

In a similar example, Elizabeth practiced techniques like giving herself safety messages, singing songs, and consciously relaxing her body while going to a café, which normally would give her severe brain fog. “*[I was] going again, and doing the same thing again, and going again. And then, by the tenth time, [my brain fog] was gone.”* And Lotte remembers her rapid improvement when she kept repeating to herself that her fatigue *“did not serve any purpose”* while engaging in activities like riding a bike or driving a car, which she had been unable to do for a while. And within a month she could do them again without problems.

Self-reassurance may sound easy, but as Paula illustrated, this often went hand in hand with an internal struggle: *“There was always another little voice, you know, saying: ‘See? You’re overdoing it. It’s just placebo. Next week you’ll crash completely.’ (…) But in the end, the positive voice kept winning: ‘No, I’m not ill. There’s nothing wrong.’”*

#### Reframing and normalising

Recognizing that words influence the subconscious brain, participants started using different words for behaviour and symptoms, to avoid illness-related words which could subconsciously contribute to the generation of symptoms. For instance, needing to rest or taking a nap, was reframed as “*my beauty sleep” (Petra)* or *“my time to chill” (Deborah).* Petra specified what this did to her: *“These are the little things that help you feel normal instead of ill.”*

Gillian reframed unpleasant symptoms during panic attacks, which happened often, by saying *“they’re having a party again in my body”*. And instead of interpreting symptoms after an activity as signs of long covid, Helen began to view them as a normal response to increasing her activity levels – much like muscle aches after going to the gym. *“Seeing my work as training softened the experience, the alarm bells didn’t go off, and it became a challenge instead.” (Helen)*

#### Ongoing effort while experiencing changes

Most participants described a gradual, step-by-step process of applying the techniques, often addressing different symptoms over time. For example, Linda started with practicing self-reassurance techniques on an old non-covid related symptom (a recurring pain in her leg, that had been bothering her for decades and would regularly wake her up at night). When this pain disappeared within two days, she felt encouraged to continue to practice with long covid symptoms. Not every participant experienced such fast changes though.

Participants described trajectories varying in how quickly they experienced improvements following engagement with mind-body techniques. For some, the sense of improvement emerged almost immediately. For example Paula, who felt stronger and fitter every day after starting with mind-body work. Others experienced a more gradual shift, or required time to explore different approaches before identifying one that resonated with them. However, even when the first relief of symptoms was noted immediately, it could take - sometimes months - of practicing mind-body techniques before they would feel (nearly) recovered. For instance Ruth describes the effort and discipline it took: “*If you don’t practise, nothing changes. (…) I didn’t finish [Breaking Free by Jan Rothney [34]] in a week (…). Each day I read one or two pages and then practised what I’d read”*. Similarly, Elizabeth describing her prolonged engagement with self-reassurance and exposure in the following way: *“It was really a matter of practising, practising, practising - 24/7 from the moment I woke up (…). You have to show your brain that everything you do is safe. And it’s not enough to just think it - you have to do it.”*

Regardless of the pace, moments of improvement were perceived as very encouraging, as proof-of-concept of the approach and were actively acknowledged and celebrated as integral to the healing process. Participants identified various milestones, such as being able to prepare tea or coffee, walking for a few minutes, making a phone call, enjoying a meal with family, watching a movie, or driving to collect their children from school. These were activities that some had been unable to for weeks or months. Petra recognized how noticing these improvements were therapeutic in itself:

> “When you can see [the small steps of improvement], write them down, and feel grateful for them, it brings a great sense of safety to the brain.”

Recovery was rarely linear; progress was often plateaued or was interrupted by setbacks like bouts of fatigue, episodes of fast heart rate and visual impairment or feelings of fatigue and brain fog.

Participants noted a growing sense of competence in applying the techniques, which occurred not only due to repeated practice but also to the easing of initial illness-related limitations as symptoms became less pronounced. Helen said it became *“a bit of a sport”* for her, and the more often she practiced, the easier it became. And Julie describes how the mind-body techniques became routine: *“I don’t need to go through all those little rituals with myself anymore - now it’s just doing it.”* This growing sense of competence was encouraging in itself and proved valuable when they encountered plateauing of progress or even setbacks, as it enabled them to draw upon the techniques they had practised to navigate these challenges more effectively.

Notably, nearly all participants undertook this process without professional (medical) support. Professional mind-body oriented support was perceived as not available or less accessible, due to high cost and lack of reimbursement through health insurance. Two participants who managed to find mind-body oriented therapists (one through an online program and the other through an in-person course) described this support as particularly helpful during periods of doubt and uncertainty about how to continue with mind-body techniques. In a few cases, mainstream healthcare providers showed openness towards mind-body approaches, which participants described as particularly encouraging and reassuring. Some participants described seeking peer support in the Dutch mind-body Facebook group.

While initial improvements were motivational, for participants it was hard to experience a halt in progress. This could make them doubt the approach or make them cease mind-body work for a while, as Nicola explains when she had her first setback: *“I thought: “the whole mind-body thing wasn’t working, it is all rubbish”, you know?”* Such episodes in the process, and the process itself, resulted for many participants in moments of evaluation and reflection: on the implementation of mind-body work and on their own circumstances.

### Evaluation of the process and reflection on factors that influence the process

Many participants spoke about moments of evaluation and reflection during their recovery journey. These reflections tended to centre around four main areas. First, some realised that some of their everyday persisting behaviours didn’t align with the mind-body approach and were therefore slowing down progress. Secondly, adopting the idea that their brain plays an important role in symptom generation, some participants felt it was important to explore underlying psychological issues that might have contributed to the persistence of their symptoms. Thirdly, many participants felt, in hindsight, that certain circumstances and personality traits at the start of their illness had made them more vulnerable to develop long covid and they identified factors that hindered their recovery process. And lastly, a number of participants recognised that they needed to change aspects of their environment - often related to work - in order to reduce the risk of future relapses.

Deborah and Maria give examples of realising that their recovery had reached a plateau due to not fully adhering to mind-body principles. Upon reflection, Deborah recognized that she continued pacing herself out of fear of overexertion: *“I stayed stuck in it, because I was dealing with fear and not fully with the mind-body approach”.* Similarly, Maria realised that maintaining a rigid daytime resting schedule gave her brain mixed messages: *“…because on the one hand I’m telling my brain and body: hey, it’s all fine. You’re safe. And on the other hand I’m saying: yes, but you still have to lie down.”* Both participants realised that these patterns of behaviour and associated thoughts might reinforce a subconscious sense of physical fragility. After increasing their activity levels and discontinuing scheduled daytime rest, they reported further improvements in their recovery.

As shown in figure 1, due to evaluation and reflection, some participants went back to their trusted mind-body sources to deepen their understanding, which led them to adjust the techniques and exercises they were using. Others, like Petra and Paula, decided to seek support from mental health professionals to explore possible underlying psychological issues. Petra noticed that she had a lot of tension in her body, and realised “*I’ve still got some work to do there. It was about things from way back.“* and Paula explained she wanted to prevent future setbacks: *“I need to deal with this in a lasting way - otherwise I’ll just end up with different symptoms again in a year”*.

With or without support from mental health professionals, many participants recognised in hindsight behavioural patterns personality traits, life circumstances and other factors that they felt had influenced the course of their illness. Frequently mentioned traits included perfectionism, people-pleasing, and a strong sense of responsibility. Elizabeth for instance became aware of *“how much pressure I put on myself. I had no idea I was a perfectionist.”* And Gillian discovered why she never wanted to ask for help: *“I felt I had to be really strong and do it all by myself, because otherwise I wouldn’t do it properly and might end up being rejected.“*

In addition, all but two participants spontaneously reported that in hindsight they came to recognise increased stress both before and during the onset of their illness, which they hadn’t experienced at the time. This included pressure at work (particularly when combined with caring responsibilities), traumatic events involving themselves or loved ones, and general anxiety linked to the covid pandemic and global political tensions. Several participants also reflected on the added strain of suffering from a new condition, noting that a lack of understanding and acknowledgment among employers and healthcare professionals - along with pressure to return to work prematurely - had further contributed to their stress.

Many participants made intentional changes in their lives due to these insights - such as switching careers entirely, cutting back on working hours, building in regular time for reflection, or taking up a new hobby. Many participants also mentioned that they actively share their insights and recovery stories with other people, hoping to help people who are now suffering from long covid, but acknowledging that not every long covid patient will be open to a mind-body perspective. Some discuss their experiences at the workplace or actively contribute to discussions in the Dutch mind-body Facebook group, others have written a book or fact sheet about their experiences or have started coaching people.

## DISCUSSION

This qualitative study aimed to explore the experiences of individuals who reported recovery from long covid while engaging with mind-body approaches. Despite variation in personal narratives, a common trajectory emerged: participants moved away from a biomedical explanatory model towards one centred on nervous system dysregulation. This shift, sometimes following initial scepticism, was often described as a turning point, sparking hope and motivation to engage in self-directed strategies. Recovery was not linear but an iterative process, involving cycles of practice, reflection (especially when progress stagnated), and adaptation. Over time, participants gained increasing confidence in their strategies, insights into contributing factors and, in many cases, made intentional life changes to support ongoing recovery. Notably, no participants reported feeling resentful about themselves or their path to recovery, although a few reflected that they would have liked to learn about mind-body work sooner after becoming ill.

Most participants navigated this process without professional support, after not making progress in mainstream healthcare. They relied instead on online mind-body focused communities, and started actively avoiding other sources of (biomedical) information that conflicted with their new understanding. To our knowledge, this is the first qualitative study to examine recovery experiences in long covid specifically through mind-body approaches, reporting common elements and themes in the recovery stories of patients. Given the qualitative and explorative design we cannot infer causality between these approaches and recovery.

Our findings echo themes reported in qualitative research on recovery from ME/CFS or chronic pain using mind-body interventions [24–26, 35–37]. These studies also report the importance of a profound shift in illness perception (from a biomedical to a mind-body perspective) in recovery, and the iterative nature of the recovery journeys. The importance of hope, and the negative effects of not being understood or acknowledged by healthcare professionals have also been described previously [21, 22, 38, 39]. On the other hand, our participants frequently emphasised their own acceptance of their current situation as a critical step, which was less evident in ME/CFS studies, but was indeed mentioned by Sarahjane Belton in the autoethnographic exploration of her own recovery experience [40]. This difference may reflect the novelty of long covid, which has prompted intense media coverage of the search for biomedical treatment, possibly resulting in prolonged hope for pharmacological treatment.

The central premise of the mind-body approach (as described by the participants) is the idea that all symptoms can be explained by nervous system dysregulation - summarised by some as ‘all symptoms are caused by the brain’. This perspective contrasts with prevailing scientific literature on the pathophysiology of long covid, which describes molecular changes and hypothesises mechanisms such as viral persistence, immune dysregulation, metabolic disturbance, and endothelial inflammation, e.g. in [8, 41–44]. None of these biomedical findings, however, offer a conclusive explanation for the full range of symptoms that has been described in long covid.

The recovery experiences of our participants challenge the biomedical model, which assumes a direct link between structural pathology and symptoms. While historically useful, this model has been criticised for oversimplifying the mind-body dualism [45] and struggles to explain persistent physical symptoms (PPS) in the absence of manifest tissue damage or physiological pathology [46]. The biopsychosocial model broadened the understanding of symptom causation but still presumes that symptoms reflect underlying pathology [47]. This dominant paradigm shapes diagnosis, care, and public expectations [48], yet does not align with our participants’ experiences.

Our participants’ accounts do however align with emerging neuroscientific perspective on symptom generation and perception, which is linked to chronic pain and other PPS [47, 49–51], and has also been suggested to play a role in long covid [14]. According to this model, any physical sensation is shaped by the brain’s predictions, which are constantly updated using bodily signals and external sensory input. Persistent symptoms can arise when the system overestimates threat, for instance after illness or a stressful situation. Although these processes take place in the subconscious, corrective experiences - such as behavioural exposure that integrates internal and external cues - may help retrain the system to recognise safety instead of threat and as a result reduce the need to produce physical warning sensations.

The mind-body techniques that were most reported by our participants (self-reassurance, exposure and reframing) fit into this neuroscientific framework, as they may act as corrective experiences, helping to recalibrate predictions and reduce symptoms. An important source of information for our participants was The Way Out by Alan Gordon and Alon Ziv and their podcast Tell Me About Your Pain, which are based on Pain Reprocessing Therapy (PRT). Symptom education and the techniques mentioned above are an important part of PRT and of Emotion Awareness and Expression Therapy (EAET) protocols. PRT and EAET share the premise that (subconscious) adaptive learning can reduce symptom activation [52] and have been shown to be effective in chronic pain and fibromyalgia [53–57]. Although evidence for their use in long covid is limited, early studies suggest potential benefit [58, 59] and larger scale prospective studies are currently being conducted or will start soon [60–62].

A major strength of this study lies in the depth and richness of the data, capturing lived experiences over time in real-world settings. The semi-structured design allowed new themes to emerge, providing insights into recovery processes that are rarely documented in clinical research. Reflexive conversations within the multidisciplinary research team contributed to a deeper understanding of the data and ensuring trustworthiness and credibility, key quality criteria for qualitative research.

However, several limitations must be acknowledged. Participants were self-identified long covid patients, without requiring formal diagnosis or positive SARS-CoV-2 testing. In addition, no strict definition of mind-body approaches was applied during recruitment, and neither was ‘recovery’ defined or quantified. These factors limit generalisability, but reflect the reality faced by clinicians encountering patients with PPS and reflect the experiences of patients, who have their own view of what recovery constitutes. Our participants described themselves as recovered, or progressing toward recovery, having transitioned from being home- or bedbound to resuming work.

Furthermore, participants were relatively highly educated and all but one white women, which limits the transferability of the findings and precludes an intersectional perspective, as experiences shaped by the interplay of sex, ethnicity, socioeconomic position and other axes of inequality are not represented.

## CONCLUSION AND FUTURE DIRECTIONS

This study provides insights into recovery experiences from long covid among individuals using mind-body approaches. Participants described a shift from a biomedical to a nervous system-based explanatory model as a turning point, fostering hope and engagement with self-directed strategies.

Their accounts align with the neuroscience of symptoms and share features with established interventions such as PRT and EAET, which have shown efficacy in chronic pain.

For clinicians who support patients with long covid, these findings suggest that acknowledging a new explanatory model for symptoms can provide a supportive clinical context that validates patients’ experiences, while allowing for behavioural experimentation. This may empower patients to look for self-management tools and facilitate recovery. For policymakers and researchers promoting a more comprehensive and inclusive understanding of long covid may reduce barriers to self-management and encourage research into novel treatment strategies. Despite these promising insights, many questions remain for future research and clinical practice.

First of all: what are the effects of mind-body interventions on long covid and how can these best be measured? Which components or elements of mind-body interventions are most effective, whether or not combined with existing therapies? And for which patient subgroups? Could certain phenotypes of long covid respond better - or worse - to these approaches? What level of public education and (professional) support is necessary for optimal outcomes? Which professional skills are necessary to offer mind-body treatments?

Addressing these questions will require development of standardised protocols for large-scale randomised controlled trials. These are needed to provide the necessary information to develop safe, evidence based and patient-centred mind-body education and interventions. Given reported negative outcomes of similar interventions by members of the long covid and ME/CFS community (but not reported in trial outcomes), it is crucial to gain insight in developing safe ways to offer mind-body treatment modalities, with careful attention to adverse events.

Future research should also explore how prevailing narratives within healthcare and society influence treatment uptake and recovery trajectories. Ultimately, integrating neuroscientific perspectives and patient experiences with symptom education and treatment programs may open new avenues for care, but this will require a shift in both clinical practice and public discourse. While we have found very positive experiences with mind-body work among our participants, it is essential to increase the safety of and negate negative experiences with these techniques, to increase the acceptability of such interventions in this contested field [25].

## Data Availability

Metadata are available at http://doi.org/10.5281/zenodo.18414706. Interview data will not be made available, given the sensitive and personal nature of this data.

## Author affiliations

All authors were affiliated with Leiden University Medical Centre, the department of Public Health and Primary Care. HV also has a GP-practice, and CD provides medical advice to the Dutch institute of asbestos victims (IAS). VB has finished her masters and is currently working in a hospital. JB is also affiliated with the institute of public administration of the faculty of governance and global affairs, Leiden University and chairs the Dutch Council for Health and Society (RVS).

## Acknowledgements

The authors are immensely grateful to the participants in this study, who donated their time and effort to share their stories of a transformative period in their lives.

We would also like to acknowledge Marjon Oomens (director of the Emovere foundation) as project manager of the Emovere project. Her energy and lived PPS experience were vital to the project. We also thank all consortium members of the Emovere project for their continuous support.

Thanks to Annemieke Ebus-Vermeer, then administrator of the Dutch mind-body Facebook group, who allowed us to recruit participants in that group.

Master student psychology Emily Jackman assisted in data collection and analysis.

## Contributors

The study was designed by CD. CD, HV and NS developed and revised the topic guide. CD did 16 interviews, a psychology master student the remaining 2. The data was coded by CD and VB. CD, VB and HV did two in-depth analysis sessions to identify themes, which were subsequently discussed with NS and JB. CD wrote the first draft and revised all subsequent drafts including the final one. All authors provided critical comments to the drafts.

The authors used Microsoft Copilot, a large language model-based tool, solely for language editing to improve clarity and grammar in the manuscript. No content, data analysis, or interpretation was generated by the AI tool. All authors reviewed and approved the final article and take full responsibility for its accuracy and integrity.

## Funding

This work is part of a citizen science and public-private partnership program. The program was initiated by a consortium of partners: the Emovere Foundation, general practitioners, health insurers, third sector organisations and Leiden University Medical Center (LUMC).

The collaboration project is co-funded by the PPP Allowance made available by Health∼Holland, Top Sector Life Sciences & Health to stimulate public-private partnerships. Stichting LSH-TKI and the Ministry of Economic Affairs and Climate Policy had no role in the design, conduct, analysis, or reporting of this study. They are not responsible for the content of this article or for any use that may be made of the findings. Grant number LSHM22007 (40-42705-98-019).

## Disclaimer

The views expressed in this publication are those of the authors and not necessarily those of the Leiden University Medical Centre, or any of the other members of the consortium.

## Competing interests

CD has developed an accredited online webinar about the role of neuroplasticity in the construction of physical sensations and symptoms. Although these seminars on neuroplasticity relate to this topic, the potential conflict was disclosed to the full research team, who jointly oversaw study design, analysis, and interpretation to prevent individual influence.

## Patient consent for publication

Not applicable

## Ethics approval

This study has been conducted according to the principles of the Declaration of Helsinki (64th WMA General Assembly, Fortaleza, Brazil, October 2013) and follows the standards of Good Research Practice. It complies with the European data protection standards (GDPR).

Ethical approval for this study was given by the department’s scientific committee (Approval ID: WSC-2023-02). This study was deemed not to fall under the Dutch law for medical research (Wmo), since it did not contain medical treatments or otherwise subject our participants to potential harmful activities (LUMC ID 22-3030).

All participants provided written informed consent prior to enrolment in the study.

## Provenance and peer review

Not commissioned; externally peer reviewed.

## Notes

### Competing Interest Statement

Christine Deurman has developed an accredited online webinar about the role of neuroplasticity in the construction of physical sensations and symptoms. Although these seminars on neuroplasticity relate to this topic, the potential conflict was disclosed to the full research team, who jointly oversaw study design, analysis, and interpretation to prevent individual influence. Other authors have no competing interests declared.

### Funding Statement

This study was part of a citizen science and public-private partnership program that was funded by funded by ZonMW Health Holland, grant number LSHM22007 (40-42705-98-019). None of the authors received payment or services from a third party for any of the submitted work.

### Author Declarations

The IRB of the Leiden University Medical Center (LUMC) waived ethical approval for this work, LUMC ID 22-3030.

