## Supplemental material for "Reclaiming health: a qualitative, explorative study of long covid recovery journeys involving mind-body approaches"

### Questionnaire with list of complaints

Which complaints did you have as a result of long covid?

| | |
|---|---|
| Fatigue | ☐ |
| Problems with concentrating | ☐ |
| Memory problems | ☐ |
| Hypersensitive to light and sound | ☐ |
| Shortness of breath | ☐ |
| Headache | ☐ |
| Feeling depressed or irritable | ☐ |
| Sleeping problems | ☐ |
| Muscle aches | ☐ |
| Dizziness | ☐ |
| Palpitations | ☐ |
| Changes in temperature | ☐ |
| Joint aches | ☐ |
| Visual impairments | ☐ |
| Abdominal complaints | ☐ |
| Chest pain | ☐ |
| (Night)sweats | ☐ |
| Abnormal (or absent) sense of smell | ☐ |
| Cough | ☐ |
| Sore throat | ☐ |
| Recurring infections | ☐ |

Other:

This list was based on the top 22 of reported symptoms according to the Dutch website C-support: https://www.c-support.nu/feiten-en-cijfers/

### Interview guide

The following demographic data will be collected at the onset of the interview: sex, age, marital status, educational background, country of birth, religious affiliation, family size, current occupation or studies.

**Idioms for complaints**

E.g. What is the word you prefer us to use for your set of complaints after your covid-infection?

**Onset of complaints**

E.g. How and when did you find out that you had a covid infection?

E.g. Can you describe your living situation when it started? (probe: uncertainty, personal issues, stress)

**Explanatory models**

E.g What did you think was going on with you? (probe: what kind of information was available and taken up, provided by whom? Friends, family, online information, social media)

OR How did you explain what you suffered from to people around you?

**Treatment**

E.g. Which treatments or strategies have you previously tried to alleviate your symptoms?

Depending on how the participant was recruited:

- Why did you respond to the request for patients to join this pilot/experiment? or
- How did you end up with this particular therapist? or
- How did you find out about mind-body therapy and what did you do to implement this yourself?

**Resources**

Which sources have helped you on your journey?

**Healing process**

What were important moments in your healing process?

If you want to explain to your friends/family etc. what helped in your recovery, what would you say?

Probe for: a gradual improvement or a distinct turning point

**Barriers and facilitators in your healing process**

Were there any barriers or setbacks in your recovery, and can you describe them?

(probe: remarks from others, medical field, social media, press, temporary worsening of symptoms)

What would you advise patients who are currently suffering from LC-19?

### Spontaneously self-reported long covid symptoms

The following symptoms were spontaneously reported by at least two participants, under “other”:

| Symptom | Number of participants who reported this |
|---|---|
| Tinnitus or hearing impairments | 6 |
| Brain fog | 4 |
| Anxiety or feelings of panic | 3 |
| PEM (post-exertional malaise) | 3 |
| A tight or numb feeling in the body, or in parts of the body | 3 |
| Feeling tense, or in fight-flight mode, "wired-but-tired" | 3 |
| Feeling overstimulated | 3 |
| POTS (postural orthostatic tachycardia syndrome) | 2 |

### Treatment modalities respondents had tried before starting a mind-body approach

| Type of treatment | Number of participants |
| --- | --- |
| physiotherapy + occupational therapy (rehabilitation care in primary care – a Dutch intervention) | 15 |
| psychological treatment | 6 |
| anti-depressants | 3 |
| second line rehabilitation care | 1 |
| anti-histamine treatment | 1 |
| speech therapy | 1 |
| *Total of mainstream therapies* | *27* |
| acupuncture | 3 |
| orthomolecular therapy | 3 |
| yoga | 2 |
| breathing exercises | 2 |
| haptotherapy | 2 |
| osteopathy | 2 |
| Chinese herbs | 1 |
| laser fysiotherapy | 1 |
| *Total of complementary and alternative treatments* | *16* |

### Sources that were consulted by the participants

Below are the sources that were mentioned by the participants. The authors do not make any recommendations or claims of efficacy about these sources and interventions.

| Category | Examples |
|---|---|
| Social media | LinkedIn.<br>Facebook: various Dutch Facebook groups about long COVID (without a mind-body focus) and the Dutch mind-body Facebook group. |
| Podcasts and webinars | “The cure for chronic pain” by Nicole Sachs (especially the episode with a Dutch woman who recovered from long-covid).<br>“Lief klein leven” (Sweet little life) by Nina Bosselaar who recovered from long-covid.<br>“Tell me about your pain” by Alan Gordon and Alon Ziv.<br>Emovere Foundation webinars.<br>“The New Mi” by Milou Pelle. |
| Audio books | “Weg van de pijn” (Away from the pain) by Saskia de Bruin<br>“The way out” by Alan Gordon & Alon Ziv |
| Books | “Weg van de pijn” (Away from the pain) by Saskia de Bruin<br>“Breaking Free” by Jan Rothney<br>“Supersnelle kennismaking met de mind-body benadering” (A superfast introduction to the mind-body approach) by Annemieke Ebus-Vermeer<br>“You are the placebo” (Joe Dispenza) |
| YouTube | The channels of Dan Buglio, Tanner Murtaugh and Jim Prussack |
| Apps | Curable |
| Websites | Stichting Emovere (The Emovere Foundation)<br>Brainproof Cognitive Movement Therapy<br>Dynamic Neural Retraining System (DNRS)<br>Howard Schubiner<br>Gupta<br>Autonomic Nervous System Reset (ANSR) |
| Journals | Interview with health care professionals |
| Programs | “Be your own medicine” by Rebecca Tolin<br>An online programme with Jim Prussack<br>The Gupta program |

### Mind-body techniques used by the participants

Most frequently reported techniques:

- Self-reassurance, often combined with the gradual resumption and build-up of activities.
- Reframing the condition, symptoms, bodily sensations, and the recovery process, by using different words to describe them.

Less frequently reported techniques, but mentioned by more than two participants:

- Meditation
- Breathing exercises
- Visualisation, either of goals or the recovery journey
- Engaging in pleasant or affectionate activities
- Learning to pause and feel - this involves consciously staying present and emotionally attuned in the moment.
- Symptom awareness and/or somatic tracking
- Exploring underlying patterns and/or emotions
- Acknowledging and/or celebrating progress
- Yoga, particularly Yoga Nidra
- Letting go of the urge to 'fix' oneself, and shifting away from an outcome-focused mindset

Other techniques (each mentioned by no more than two participants):

- The Curable app
- The Gupta Programme
- Cold water immersion (ice water baths)
- Infrared exercise therapy
- Acceptance and Commitment Therapy (ACT)
- Chinese herbal medicine
- Eye Movement Desensitisation and Reprocessing (EMDR)
- Massage therapy
- Mindfulness
- Vagal nerve piercing
- Pacing
- Writing or journalling
- Stress-focused therapy
- Trauma Releasing Exercises (TRE)
- Regular self-reflection
